# ReCOGnAIze app to detect mild cognitive impairment and vascular cognitive impairment

**DOI:** 10.1101/2025.05.10.25327352

**Authors:** Adnan Azam Mohammed, Ashwati Vipin, Leow Yi Jin, Eliana Setiabudi, Farid Tan, Hitesh Agarwal, Kai Xin Liau, Pricilia Tanoto, Shan Yao Liew, Bocheng Qiu, Gurveen Kaur Sandhu, Jia Dong James Wang, Kiirtaara Aravindhan, Nagaendran Kandiah

## Abstract

**INTRODUCTION:** Mild Cognitive Impairment (MCI) is underdiagnosed due to subtle symptoms. Vascular Cognitive Impairment (VCI), the second leading cause of cognitive impairment, remains underdiagnosed due to varying non-amnestic manifestations. We aimed to develop and validate ReCOGnAIze, a gamified, interpretable digital app to detect MCI and VCI.

**METHODS:** A multi-phase, cross sectional study in an Asian community cohort with development phase (n=200) and validation with 235 independent participants having comprehensive neuroimaging and neuropsychological data.

**RESULTS:** In 235 subjects, ReCOGnAIze composite score accurately distinguished MCI from cognitively normal individuals (AUC = 0.90), outperforming the Montreal Cognitive Assessment (AUC = 0.70). For VCI detection, it achieved strong performance (n = 155; Average AUC = 0.85), identifying biomarkers of VCI; response time variability, task efficiency, consistent with known cerebrovascular disease impairments.

**DISCUSSION:** ReCOGnAIze is a scalable, explainable AI tool that accurately detects MCI and VCI, with interpretable clinical insights.

## 1. BACKGROUND

Mild cognitive impairment (MCI) is widely recognized as a prodromal stage of dementia, with a significant 10–15% annual risk of conversion from MCI to dementia [1,2]. Given the global burden of dementia—currently affecting over 57 million individuals, projected to reach 150 million by 2050, the societal and economic impacts are profound [3]. With emerging pharmacological and non-pharmacological interventions demonstrating potential to delay dementia onset, even a modest delay of five years could reduce its prevalence by nearly 50%, underscoring the critical importance of early identification and intervention in MCI [4,5].

Vascular cognitive impairment (VCI), the second most common cause of cognitive decline globally, accounts for more than 20 million dementia cases [6]. Although there have been substantial advancements in pharmacotherapies for Alzheimer’s Disease (AD) with monoclonal antibodies targeting amyloid-β, these treatments may be less effective in VCI and carry increased risk of cerebral haemorrhage [7]. By contrast, VCI may respond to specific lifestyle interventions, and intensive management of vascular risk factors, such as hypertension, diabetes, and hyperlipidaemia [8]. The landmark SPRINT MIND study demonstrated that intensive blood pressure control could significantly reduce progression of cognitive impairment [9]. This highlights the importance of detecting and treating VCI accurately and early, to reduce the risk of future dementia and stroke.

The burden of VCI is particularly high in Asian, African, and Hispanic populations, where elevated rates of hypertension, ischemic stroke, and diabetes drive cognitive decline [10–16]. A multi-country study across nine Asian cities highlighted the widespread prevalence of cerebrovascular disease and its detrimental impact on cognition [17]. Despite this, conventional cognitive screening tools such as the Mini-Mental State Examination (MMSE) and Montreal Cognitive Assessment (MoCA) are optimized for memory deficits, limiting their utility in detecting hallmark VCI features, including slowed processing speed [18]. While comprehensive neuropsychological batteries for VCI exist, they are time-consuming and impractical for large-scale screening [19,20].

Digital cognitive assessments offer a promising solution to cognitive screening with self-administered, remote, repeatable, and automatically scored objective cognitive tests [21]. However, many are not specific for VCI. Commercially available platforms such as the CANTAB, Cognigram, and Neurotrack have primarily been validated for MCI of the AD type, rather than VCI [20]. Digital tools have rarely incorporated cerebrovascular markers in their development and validation, limiting their capability in differentiating between AD-related MCI and VCI [22]. Hence, there is an urgent need for screening tools to detect both MCI and VCI, given the rising burden of vascular risk factors, sedentary lifestyle and genetic predispositions making populations vulnerable to VCI [22].

To address this critical gap, we designed and developed the ReCOGnAIze App—a gamified digital cognitive assessment, for detecting MCI and VCI, validated in a community cohort having a high cerebrovascular burden. Gamified cognitive tasks with explainable AI models hold the potential to significantly enhance the early identification and differentiation of MCI and VCI, laying the groundwork for precise, scalable screening [8,9].

## 2. METHODS

ReCOGnAIze was designed, developed, and validated in two phases using data from participants in the Biomarker and Cognitive Study, Singapore (BIOCIS) [23].

### 2.1 Study Population

BIOCIS participants were classified as Cognitively Normal (CN) or as having MCI based on Peterson’s criteria and National Institute on Aging-Alzheimer’s Association guidelines after completion of a validated neuropsychological test battery [1,24]. Participants with MCI had a score of more than 4 on the Subjective Memory Questionnaire, had cognitive deficits 1.5 standard deviation (SD) below age matched norms, a global clinical dementia rating (CDR) score of 0.5-1, and had no functional deficits [25]. CN subjects had a global CDR of 0 and no significant neurocognitive deficits. Further methodological details are available in the BIOCIS protocol paper [23,26].

VCI was defined as per the Diagnostic and Statistical Manual of Mental Disorders, Fifth Edition criteria and included cognitively impaired (either subjective or objective) individuals with neuroimaging evidence of vascular etiology identified by moderate-to-high levels of WMH on Brain-MRI (modified Fazekas score of 5 to 12). Non-vascular cognitively impaired (NVCI) included cognitively impaired (either subjective or objective) individuals with low levels of WMH on Brain-MRI (modified Fazekas score of 0 to 4) [6,11,27].

Two independent cohorts were used for development and validation. The development cohort comprised 200 subjects having demographic, neuropsychological and MRI data. The validation cohort included 235 subjects having demographic, neuropsychological and MRI data. All participants provided informed consent, and the study was approved by the institutional ethics board (IRB no: IRB-2021-1036).

### 2.2 Neuropsychological assessments and demographic data

Briefly, demographic variables included age, sex, ethnicity, information about vascular risk factors: Diabetes Mellitus (DM), Hypertension (HTN), and Hyperlipidaemia (HLD) as defined by the clinical guidelines from the Ministry of Health, Singapore [28–30].

Neuropsychological measures included MoCA, Visual Cognitive Assessment Test (VCAT) [31], and composites Z scores of five cognitive domains: episodic memory (Rey Auditory Verbal Learning Test [RAVLT] delayed, Wechsler Memory Scale Fourth Edition [WMS-IV] Logical Story delayed, Rey-Osterrieth Complex Figure Test [RCFT] delayed), executive function (Colour Trails [CT] 2, Wechsler Adult Intelligence Scale Fourth Version [WAIS-IV] Digit span backward), processing speed (Trail Making Test [TMT] B, CT1, Symbol Digit Modalities Test [SDMT]), visuospatial (RCFT copy and WAIS-IV Block Design), and language domains (semantic fluency animals MoCA and semantic fluency vegetables VCAT) [32–39]. Participants underwent pen and paper neuropsychological testing facilitated by a trained researcher. Information about behaviour was collected using the mild behavioural impairment checklist (MBI-C) [40].

### 2.3 Neuroimaging data

Brain imaging was performed using the Siemens 3-Tesla Prisma MRI. The T1-weighted Magnetization Prepared Rapid Gradient Echo and Fluid Attenuated Inversion Recovery (FLAIR) sequences scan obtained was pre-processed using the Computational Anatomy Toolbox in the Statistical Parametric Mapping (SPM12) toolbox on MATLAB 2022b [41]. Measures of WMH through the Fazekas Rating and other measures of cerebral small vessel disease using the Staals’ criteria, were quantified and obtained by trained visual raters [42,43].

### 2.4 Phase 1: Data-Guided Development of the App

#### 2.4.1 Cognitive-behavioural profiles associated with MCI and VCI

BIOCIS data from a sub-group of 200 subjects (Development Cohort) was analyzed [23] and multivariate analyses compared cognitive-behavioral profiles with MCI groups with moderate-to-high WMH and low WMH, controlling for age, education, APOE4 carrier status. MCI individuals with moderate-to-high WMH demonstrated significantly greater impairment in executive function, processing speed, and impulse control, with trending decreased motivation, measured by the mild behavioural impairment checklist (MBIC) [40], further shown in **Figure 1**. These findings were important in developing a tool that can not only detect MCI, but also differentiate VCI. These were incorporated in cognitive game development.

**Figure 1.**
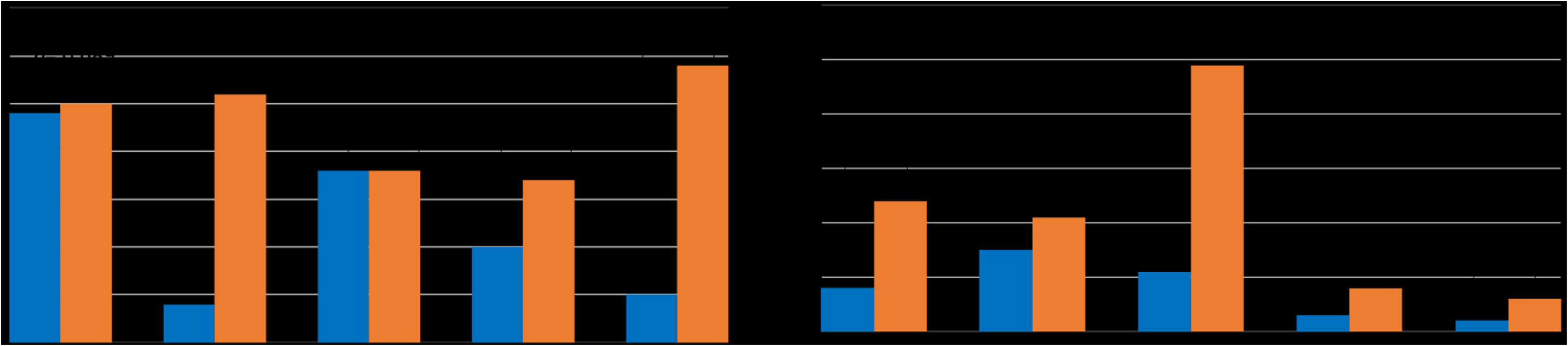
Cognitive-behavioral profile differences between MCI participants with and without VCI. Multivariate analysis of the development cohort (n = 200) comparing MCI participants with vascular cognitive impairment (VCI) to those without VCI (non-VCI) revealed significantly greater impairments in executive function, processing speed, and impulse control among the VCI group, after adjusting for age, education, and APOE4 carrier status. A trend toward reduced motivation was also observed, as measured by the Mild Behavioural Impairment Checklist (MBIC). These domain-specific deficits informed the selection and design of gamified cognitive tasks in the ReCOGnAIze app.

#### 2.4.2 Development of gamified cognitive tasks

Based on the cognitive-behavioural characterization, 4 gamified cognitive tasks were designed and developed as shown in **Figure 2**. The tasks comprised of: Symbol Matching – based on the Digit Symbol Substitution Test intended to primarily assess Processing Speed; Trail Making – adapted version of the Trail Making Test intended to primarily assess Executive Function; Airplane Game – go/no-go paradigm intended to primarily assess Attention and Impulse Control; and Grocery Shopping – a Memory and Processing Speed assessment with increasing difficulty levels intended to assess memory, processing speed, impulse control and motivation [37,38]. The cognitive tasks are further described below:

**Figure 2.**
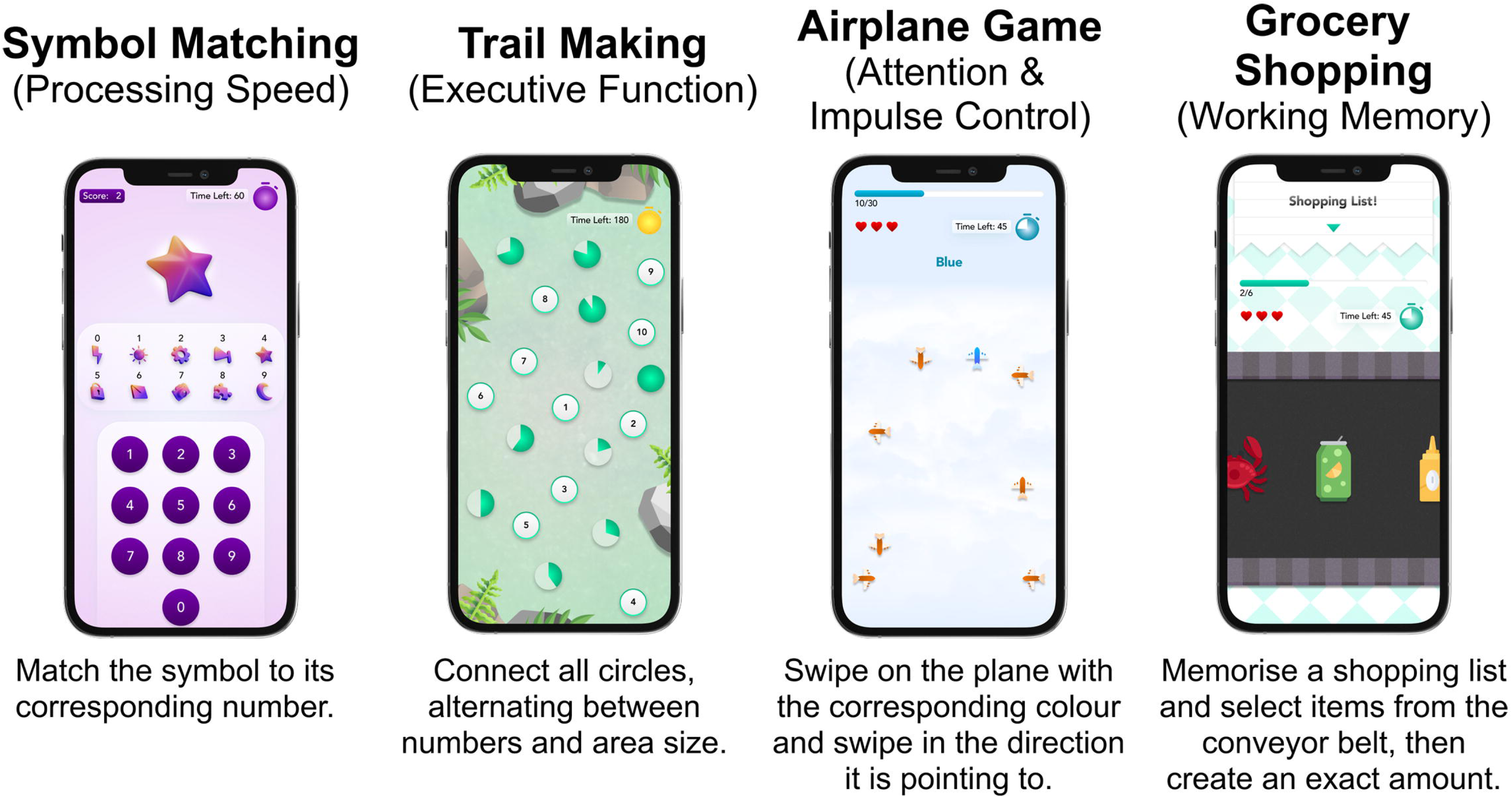
ReCOGnAIze Digital Cognitive Assessments and Their Targeted Domains. Four gamified, each designed to evaluate a distinct cognitive domain. Symbol Matching evaluates processing speed by requiring users to match abstract symbols to corresponding numbers. Trail Making evaluates executive function through an alternating attention task where users connect numbers within a green zone. Airplane Game evaluates attention and impulse control by asking users to swipe in the direction of a colored airplane while ignoring distractors. Grocery Shopping evaluates working memory and processing speed by memorizing a shopping list and selecting items from a conveyer belt in a limited time.

##### 2.4.2.1 Symbol Matching (Processing Speed)

A task requiring users to match symbols to their corresponding numbers within 1 minute with continuous randomization. This game traditionally measures information processing speed, which is commonly affected in patients with hallmark of subcortical ischemic VCI, where small vessel disease disrupts frontal-subcortical circuits. The game was chosen based on its strong literature backing as a sensitive marker of processing speed deficits [6,44].

Usability testing ensured that task complexity matched participants’ cognitive capabilities, refining the interface to optimize engagement and response accuracy. The game was designed with simple, intuitive controls and clear visual cues to ensure ease of interaction for users with varying levels of cognitive function.

##### 2.4.2.2 Trail Making (Executive Function)

An adaptation of the traditional trail-making test where users must connect all circles within 3 minutes, alternating between numbers and shapes, making it language-neutral. This task assesses executive function, cognitive flexibility, and set-shifting abilities, which are particularly vulnerable to frontal-subcortical circuit disruption in vascular cognitive impairment (VCI) [18,45]. Literature supports trail-making tests as a reliable indicator of executive dysfunction, making it a key inclusion in our app [35,37,46,47]. Focus groups provided feedback on time constraints, ensuring task difficulty was appropriate for older adults with varying cognitive abilities. The interface was designed to be highly responsive with clear step-by-step guidance, making it accessible even to individuals with mild motor impairments.

##### 2.4.2.3 Airplane Game (Attention and Impulse Control)

A go/no-go paradigm where users focus on the plane’s color and swipe in its pointing direction. This game measures response inhibition and impulse control [48,49]. Impulse dyscontrol is a significant behavioral manifestation in patients with high cerebrovascular burden. Selection of this game was further supported based on evidence supporting go/no-go paradigms in detecting inhibitory control deficits [50,51]. Usability testing refined task instructions and response timing to enhance accessibility and minimize errors unrelated to cognitive impairment. The game features a user-friendly design with large, easy-to-identify visual elements and minimal text requirements, making it suitable for users with varying literacy levels and cognitive abilities.

##### 2.4.2.4 Grocery Shopping (Memory)

A task requiring users to memorize a shopping list, select items from a conveyor belt, and prepare exact change. This game evaluates memory and attention. Selection of this task was supported by its ecological validity in assessing real-world memory function [52,53]. Focus groups provided feedback on task realism, improving user engagement and refining difficulty settings to balance challenge and feasibility. The game was designed with a visually engaging and interactive format to simulate a real-world shopping experience, ensuring it remains intuitive and enjoyable while accurately assessing memory function.

#### 2.4.3 Integration of gamified cognitive tasks into a structured guided assessment app

To ensure seamless and intuitive user experience, individual cognitive games were integrated into a structured framework with step-by-step instructions for each task, ensuring understanding and completion without external assistance. Each game was accompanied by guided tutorials, visual cues, text-based prompts, and interactive practice trials to familiarize users with the mechanics before starting the assessment.

The assessment followed a **Learning -> Trial -> Assess** framework. Users first engage in a guided learning phase where they are introduced to the task through step-by-step explanations and demonstrations. This is followed by a trial phase, allowing users to practice in a low-stakes environment with real-time feedback to build confidence and familiarity. Finally, the test phase objectively measures cognitive performance under standardized conditions, ensuring valid and reliable results.

User feedback from pilot testing and focus groups informed iterative refinements in the instruction delivery and task order. Adjustments were made to optimize user comprehension, minimize errors due to misunderstanding, and enhance the overall experience of taking the assessment. Through these enhancements, the ReCOGnAIze app was transformed into a comprehensive, structured cognitive assessment tool that is both scientifically robust and highly accessible.

### 2.5 Phase 2: Scoring and Validation of the App

#### 2.5.1 Study Population (Validation Cohort)

An independent cohort of 235 participants was recruited from the BIOCIS study for testing and validation of ReCOGnAIze for detection of MCI and differentiation of VCI from NVCI [23]. Participants underwent the assessment on the ReCOGnAIze web application. Data collected was further processed as detailed below.

#### 2.5.2 Composite Cognitive Performance Scores

To validate ReCOGnAIze’s cognitive assessment ability, we calculated a normalized, equally-weighted composite score *Recognaize*_*total* to reflect global cognition.

Each of the four games in ReCOGnAIze was used to generate an individual score based on accuracy of task completion and response time, computed as:

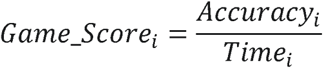

These were normalized using Min-Max scaling, mapping each score to a range between 0 and 5:

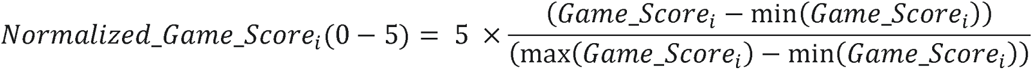

where *Normalized_Game_Score_1_* is the normalized score for game *i*, and min(*Game_Score_1_*)and max(*Game_Score_1_*) represent the minimum and maximum raw scores observed in the sample for that game.

The ReCOGnAIze Composite Score was then calculated as the summation of the four normalized game scores:

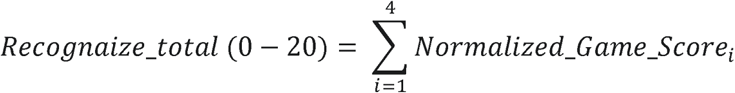

#### 2.5.3 Feature Engineering and Selection for VCI

Feature engineering was done to extract granular game features of attention, processing speed and executive function: time-based, accuracy, performance consistency, and efficiency, to enable a deeper investigation into the pathobiology of VCI.

Time-based measures capture response speed and variability, reflecting cognitive efficiency and flexibility. Central tendency statistics (mean, median, mode) determined typical response speed, while range measures (minimum, maximum) highlighted performance fluctuations. Variability metrics (standard deviation, interquartile range (IQR), CV) assessed response stability, as greater variability could indicate cognitive decline. Percentile measures (25th, 75th) indicated shifts in response distribution, and time trend analyses examined changes over the task, revealing fatigue, learning effects, or attentional lapses.

Accuracy measures were used to assess task performance and error patterns. Accuracy changes between the first and second halves of tasks could identify cognitive fatigue or adaptation. Performance consistency reflected cognitive stability and resilience, with sustained attention measures like longest streak of correct or incorrect responses. Efficiency measures integrated performance indices and efficiency scores to identify compensation strategies.

To determine the most predictive, interpretable digital cognitive features for VCI, we applied a three-stage pipeline. First, we computed SHapley Additive Explanations (SHAP) values from a Random Forest model and retained the top 30 most important features [54]. Second, we applied Elastic Net regularization to select robust predictors while accounting for multicollinearity between features [55]. Third, we performed Recursive Feature Elimination with cross-validation (RFECV) using logistic regression to identify the minimal subset of features with maximal predictive performance for VCI [56].

#### 2.5.4 Vascular Cognitive Scores

Selected features were combined with demographic and vascular risk factors associated with VCI — Age, Diabetes Mellitus (DM), Hypertension (HTN), and Hyperlipidaemia (HLD) [57]—to construct Vascular Cognitive Scores (VCS) using both rule-based and ML approaches.

We computed *Recognaize_VCS_Composite_*, a normalized, equally-weighted mean of selected cognitive features and risk variables:

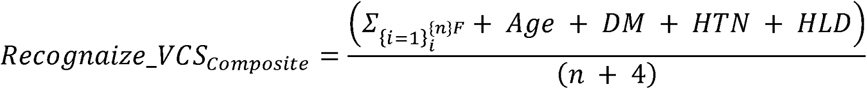

where *F_n_*represents the selected digital cognitive features.

ML classifiers: Random Forest, XGBoost, LightGBM, and CatBoost were also trained to detect VCI, yielding ML-derived VCS which were validated with 5 fold cross validation (CV) in detecting VCI. Mathematically:

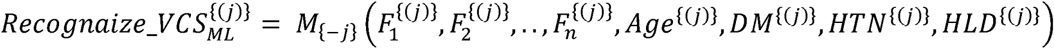

where *M*_{-*j*}_ denotes the ML model trained on all data excluding fold *j*, and superscript (*j*) indicates the data of the held-out fold. For unbiased and generalizable models, all ML-based VCS were generated using out-of-fold (OOF) predictions from a 5-fold Cross-Validation (CV) procedure. Each model was trained on 80% of the dataset and predict VCI for the held-out 20%. This process was repeated across all five folds such that each subject’s VCS was obtained from a model that had not been trained on their data. This approach ensures that no subject’s score was influenced by a model trained on their own data, reducing overfitting risk and improving downstream analyses’ robustness.

### 2.6 Statistical Methods

All statistical analyses were conducted using Python (v3.9, Scipy, Scikit-learn, CatBoost, LightGBM, XGBoost, SHAP, ElasticNet, RFECV) [54–56,58–63].

#### 2.6.1 Participant Characteristics

Baseline characteristics of the study cohort were summarized using means and SD for continuous variables. Categorical variables were presented as percentages. Group comparisons between CN and MCI individuals, and between VCI and NVCI individuals, were performed using independent t-tests for continuous variables and chi-squared tests for categorical variables.

For cognitive outcome measures, additional age- and education-adjusted comparisons were performed using analysis of covariance (ANCOVA), with diagnostic group as the between-subjects factor and age and years of education as covariates. Adjusted means ± standard errors (SE), *p* values, and partial eta squared (eta²) effect sizes were reported. Cohen’s *d* was calculated for unadjusted comparisons to estimate effect sizes. All *p* values were two tailed, and a threshold of *p*<0.05 was considered statistically significant.

#### 2.6.2 Differentiating MCI from CN

The ability of the *Recognaize_total* composite score to differentiate MCI from CN was assessed using receiver operator characteristic (ROC) curves, with area under the curve (AUC), 95% confidence interval, sensitivity, specificity reported. To benchmark diagnostic performance, we compared the *Recognaize_total* Composite Score against established MCI screening tools: MoCA and VCAT [31,64]. The optimal cut-off for MCI detection was determined using Youden’s J statistic [65].

#### 2.6.3 Differentiating VCI from NVCI

The ability of the different VCS scores to differentiate VCI from NVCI was assessed using 5-fold CV average AUC with accuracy, sensitivity, specificity and F1-score (for ML models) reported.

## 3. RESULTS

### 3.1 Participant Characteristics

As shown in **Table 1**, from 235 total, MCI participants (n = 108; mean age, 65.0 years [SD 9.0]) were significantly older than CN (n = 127; 58.2 years [SD 9.6]; *p* < .001, *d* = 0.74). MCI participants also had fewer years of education (13.9 [SD 3.4] vs 15.1 [SD 2.8]; *p* = .004, *d* = 0.39). In age- and education-adjusted analyses, MCI participants had significantly lower MoCA scores (adjusted mean, 25.16 [SE 0.24] vs 26.59 [SE 0.22]; *p* < .001; partial eta² = 0.07) and VCAT scores (26.31 [SE 0.24] vs 27.20 [SE 0.22]; *p* = .009; partial eta² = 0.03), and a large group difference in executive function (–0.67 [SE 0.06] vs –0.00 [SE 0.06]; *p* < .001; partial eta² = 0.19) (Table 1).The *ReCOGnAIze_total* composite showed the largest group difference (adjusted mean, 7.72 [SE 0.17] vs 10.47 [SE 0.16] ; *p* < .001; partial eta² = 0.36), outperforming MoCA (eta² = 0.07) and VCAT (eta ² = 0.03). ReCOGnAIze’s individual game scores also significantly differed between groups with large effect sizes: Trail Making (eta² = 0.18), Airplane Game (eta² = 0.16), Grocery Shopping (eta² = 0.18), and Symbol Matching (eta² = 0.13) (Table 1).

**Table 1.**
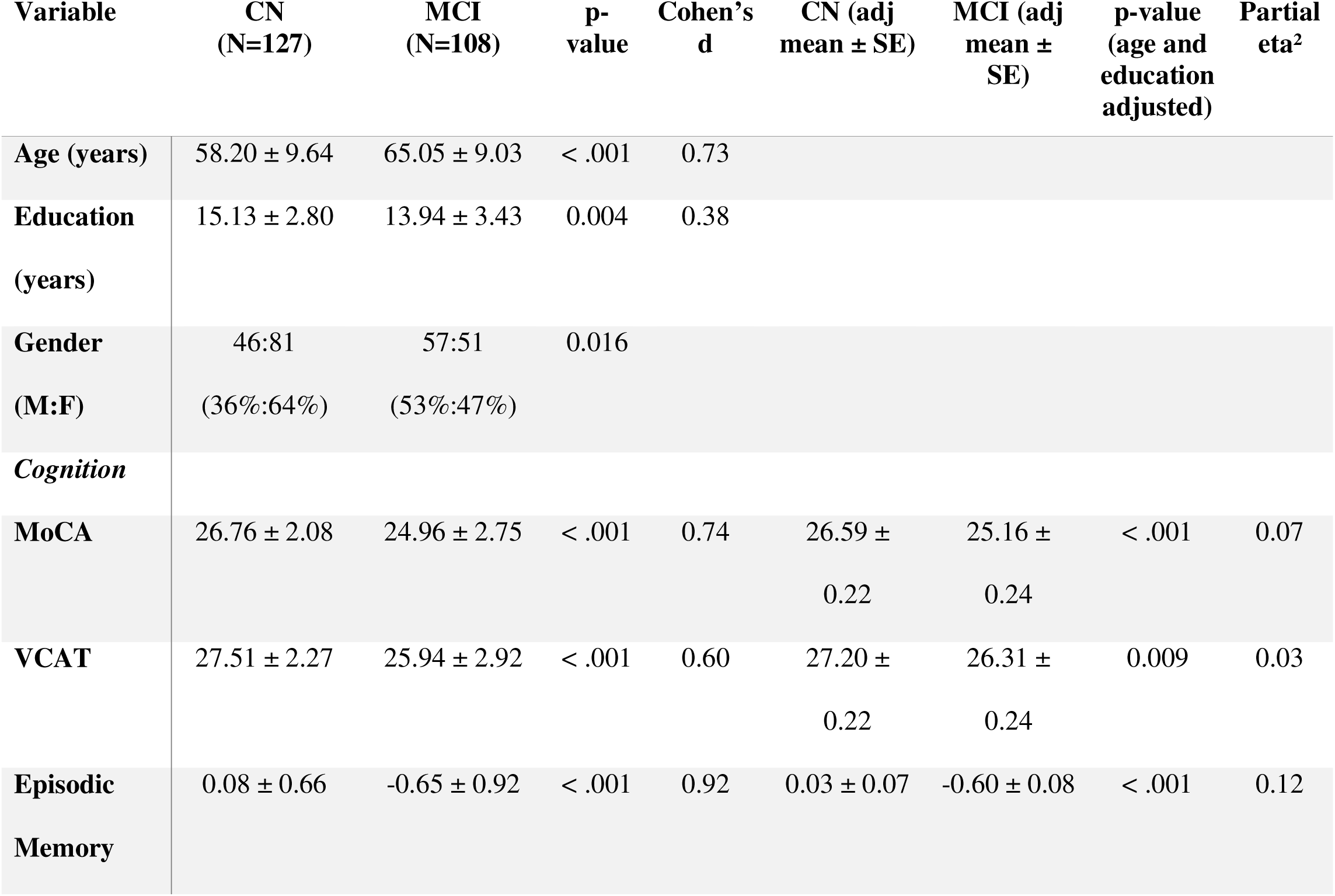
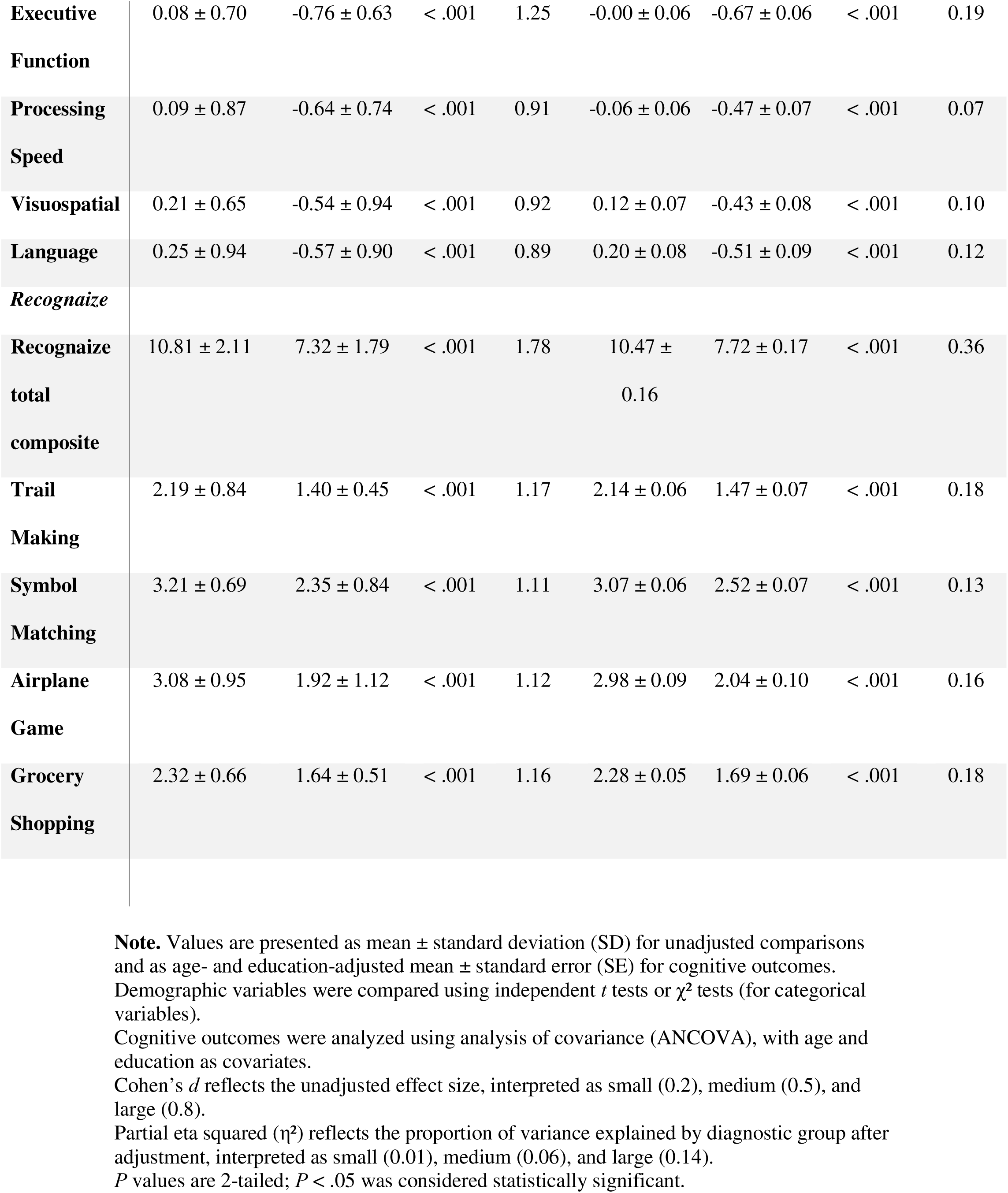
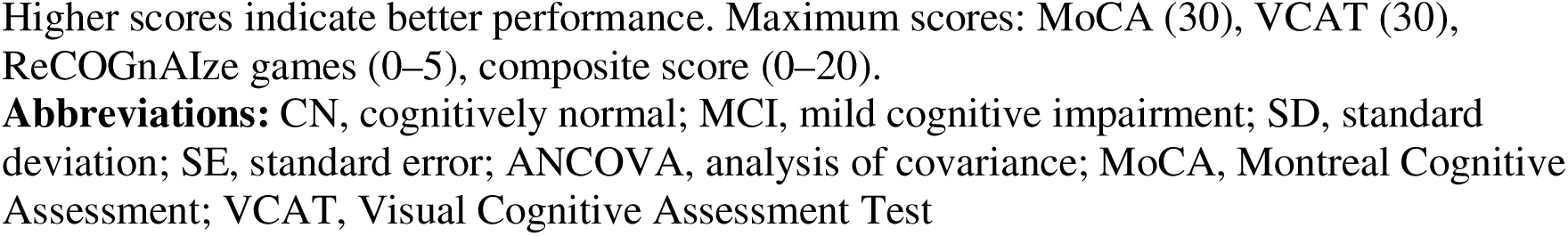
Comparison of Demographic Characteristics and Cognitive Scores Between Cognitively Normal and Mild Cognitive Impairment Groups.

### 3.2 Detection of Mild Cognitive Impairment

As shown in **Figure 3**, ROC analyses comparing MCI and CN demonstrated that ***Recognaize_total* composite score achieved** the highest diagnostic accuracy (AUC = 0.90, 95% CI 0.87–0.94) with a sensitivity of 0.85 and specificity of 0.84 at the optimal threshold detailed in **Table 2**. This outperformed traditional tools, **MoCA** (AUC = 0.70, 95% CI 0.63– 0.77) and **VCAT** (AUC = 0.66, 95% CI 0.59–0.73) as shown in **Figure 3, Panel A**.

**Figure 3.**
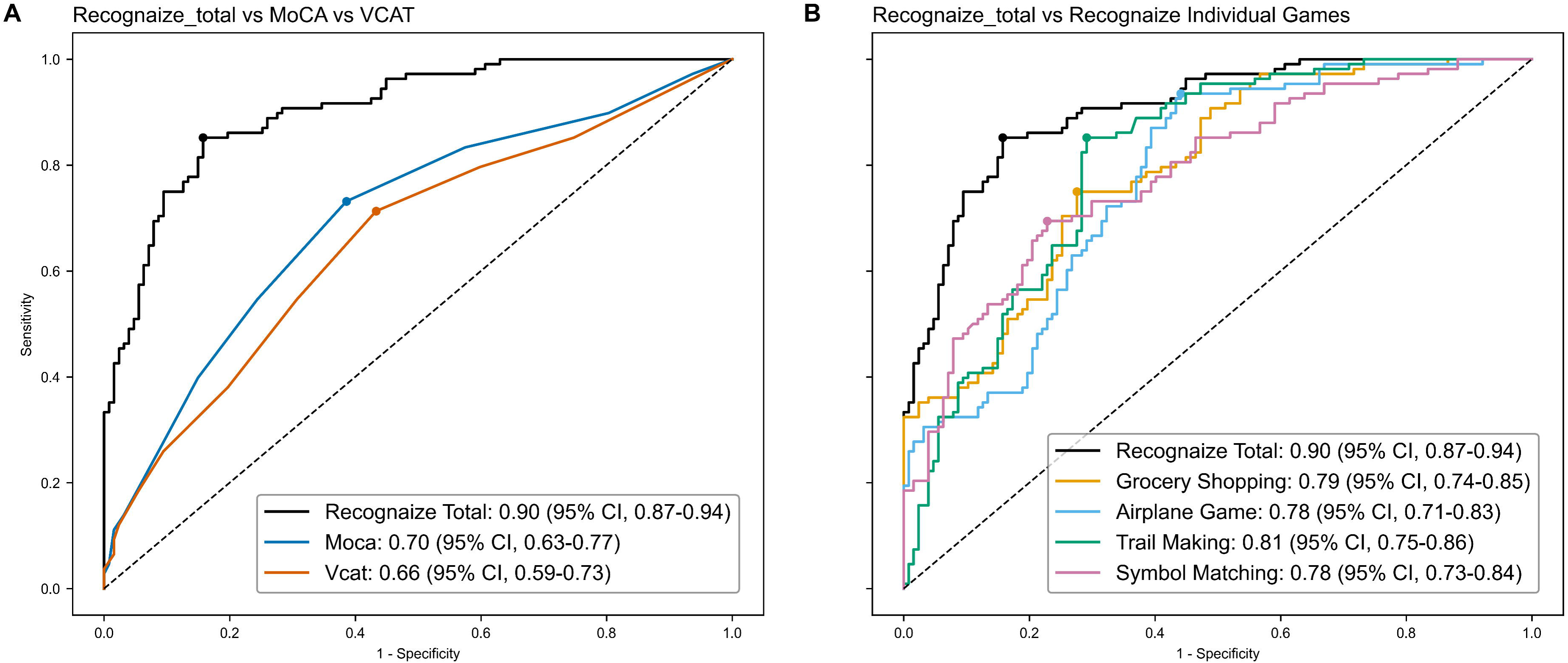
Diagnostic accuracy of ReCOGnAIze composite score, and individual game scores, compared to traditional tests, in detecting MCI. Panel A shows ROC curves for ReCOGnAIze total score, Montreal Cognitive Assessment (MoCA), and Visual Cognitive Assessment Test (VCAT) in distinguishing individuals with mild cognitive impairment (MCI) from cognitively normal (CN) individuals. Panel B displays ROC curves for the ReCOGnAIze total score and individual game scores: Trail Making, Grocery Shopping, Symbol Matching, and Airplane Game. Circles indicate optimal operating points based on the Youden index. Area under the curve (AUC) values with 95% confidence intervals (CIs) are reported in the legend. ReCOGnAIze outperformed traditional tools and demonstrated strong discriminative ability across both total and individual game scores.

**Table 2.**
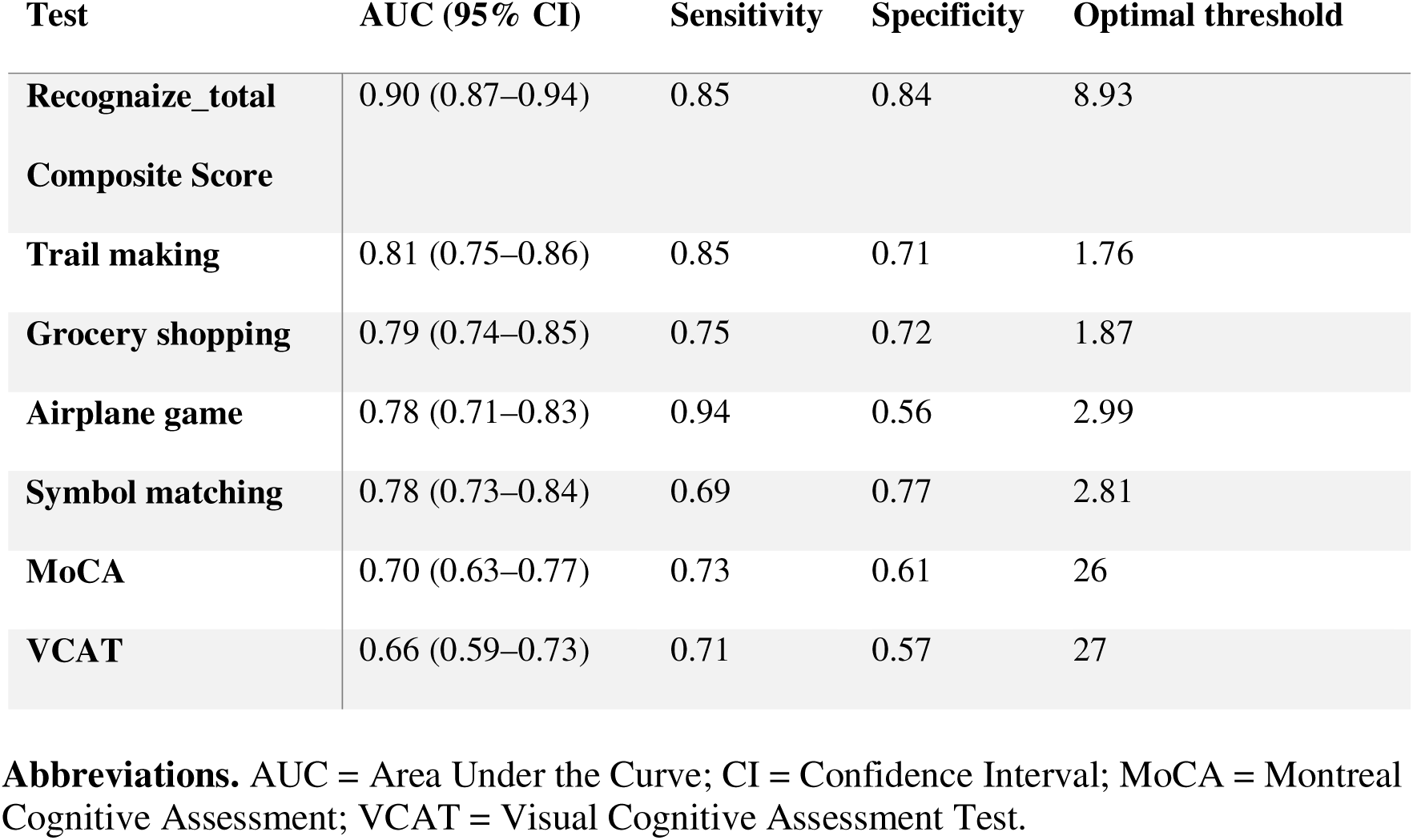
Diagnostic performance of ReCOGnAIze, its subtests, the Montreal Cognitive Assessment and the Visual Cognitive Assessment Test for detecting Mild Cognitive Impairment with area under the curve, sensitivity, specificity, and optimal threshold reported.

Figure 3, **Panel B** shows that individual ReCOGnAIze games also demonstrated robust classification performance: **Trail Making** (AUC = 0.81, 95% CI 0.75–0.86), **Grocery Shopping** (AUC = 0.79, 95% CI 0.74–0.85), **Symbol Matching** (AUC = 0.78, 95% CI 0.73–0.84), and **Airplane Game** (AUC = 0.78, 95% CI 0.71–0.83). AUCs, optimal thresholds, specificity and sensitivity for all tests are provided in **Table 2**. These findings support the validity of ReCOGnAIze and its component tasks in detecting MCI.

### 3.3 Detection of Vascular Cognitive Impairment

As shown in **Table 3**, participants with VCI (n = 75; mean age, 67.1 years [SD 7.6]) were significantly older than those without VCI (n = 79; 60.6 years [SD 9.0]; *p* < .001, *d* = 0.78). Hyperlipidaemia was significantly more common in the VCI group. White matter disease burden was higher in the VCI group, with higher Fazekas Total scores and WMH volumes. Machine learning–based Vascular Cognitive Scores showed the strongest separation between groups even after adjusting for age: CatBoost-VCS (adjusted mean, 0.62 [SE 0.02] vs 0.39 [SE 0.02]; *p* < .001; eta² = 0.22) and XGBoost-VCS (0.66 [SE 0.04] vs 0.29 [SE 0.04]; *p* < .001; eta² = 0.22), outperforming MoCA and VCAT.

**Table 3.** Comparison of Demographic Characteristics, Imaging Markers, Cognitive Scores and ML-Derived Vascular Cognitive Scores between VCI and NVCI sub-groups.

| Variable | VCI<br>(N=75) | NVCi<br>(N=79) | p-value | Cohen's d | VCI (adj<br>mean ± SE) | NVCi (adj<br>mean ± SE) | p-value<br>(age-<br>adjusted) | Partial<br>eta<sup>2</sup> |
| --- | --- | --- | --- | --- | --- | --- | --- | --- |
| Age (years) | 67.08 ± 7.63 | 60.55 ± 9.03 | < .001 | 0.78 |  |  |  |  |
| Education<br>(years) | 13.96 ± 3.24 | 14.35 ± 3.21 | 0.450 | 0.12 |  |  |  |  |
| Diabetes<br>Mellitus | 17 (22.7%) | 15 (19.0%) | 0.234 |  |  |  |  |  |
| Hypertension | 28 (37.3%) | 21 (26.6%) | 0.137 |  |  |  |  |  |
| Hyperlipidemia | 45 (60.0%) | 28 (35.4%) | 0.003 |  |  |  |  |  |
| *Cognition* |  |  |  |  |  |  |  |  |
| **MoCA** | 24.93 ± 2.31 | 25.94 ± 2.70 | 0.015 | 0.40 | 25.13 ± 0.30 | 25.75 ± 0.29 | 0.149 | 0.01 |
| **VCAT** | 26.12 ± 2.55 | 26.81 ± 2.90 | 0.119 | 0.25 | 26.42 ± 0.32 | 26.53 ± 0.31 | 0.818 | 0.00 |
| **Episodic**<br>**Memory** | -0.42 ± 0.83 | -0.41 ± 0.96 | 0.909 | 0.02 | -0.35 ± 0.11 | -0.48 ± 0.10 | 0.392 | 0.00 |
| **Executive**<br>**Function** | -0.58 ± 0.75 | -0.35 ± 0.76 | 0.058 | 0.31 | -0.48 ± 0.09 | -0.45 ± 0.08 | 0.840 | 0.00 |
| **Processing**<br>**Speed** | -0.59 ± 0.81 | -0.25 ± 0.80 | 0.010 | 0.42 | -0.43 ± 0.08 | -0.41 ± 0.08 | 0.881 | 0.00 |
| **Visuospatial** | -0.47 ± 0.97 | -0.11 ± 0.83 | 0.015 | 0.40 | -0.36 ± 0.10 | -0.21 ± 0.10 | 0.334 | 0.01 |
| **Language** | -0.27 ± 0.91 | -0.19 ± 0.94 | 0.567 | 0.09 | -0.18 ± 0.11 | -0.27 ± 0.10 | 0.566 | 0.00 |
| *Imaging* |  |  |  |  |  |  |  |  |
| **Modified**<br>**Fazekas** | 7.43 ± 2.13 | 2.38 ± 1.48 | < .001 | 2.76 | 7.16 ± 0.20 | 2.63 ± 0.20 | < .001 | 0.61 |
| **WMH**<br>**(CAT12)** | 4.44 ± 3.43 | 1.28 ± 0.77 | < .001 | 1.27 | 4.17 ± 0.28 | 1.54 ± 0.28 | < .001 | 0.21 |
| *Recognize*<br>*VCS models* |  |  |  |  |  |  |  |  |
| **Recognize_**<br>**vcs_randomf**<br>**orest** | 0.61 ± 0.19 | 0.37 ± 0.18 | < .001 | 1.26 | 0.57 ± 0.02 | 0.41 ± 0.02 | < .001 | 0.18 |
| **Recognaize_**<br>**vs_catboost** | 0.66 ± 0.23 | 0.35 ± 0.23 | < .001 | 1.36 | 0.62 ± 0.02 | 0.39 ± 0.02 | < .001 | 0.22 |
| **Recognaize_**<br>**vcs_xgboost** | 0.70 ± 0.36 | 0.25 ± 0.32 | < .001 | 1.32 | 0.66 ± 0.04 | 0.29 ± 0.04 | < .001 | 0.22 |
| **Recognaize_**<br>**vcs_lightgb**<br>**m** | 0.70 ± 0.31 | 0.29 ± 0.31 | < .001 | 1.34 | 0.66 ± 0.04 | 0.32 ± 0.03 | < .001 | 0.23 |

**Note.** Values are presented as mean ± standard deviation (SD) for unadjusted comparisons and as age- and education-adjusted mean ± standard error (SE) for cognitive outcomes.
Demographic variables were compared using independent *t* tests or $\chi^2$ tests (for categorical variables). Cognitive outcomes and vascular cognitive scores (VCS) were analyzed using analysis of covariance (ANCOVA), with age as a covariate.
Higher cognitive scores indicate better performance. Higher Fazekas Score and higher WMH volumes indicate higher neuroimaging presence of WMH. Higher VCS scores indicate higher probability of VCI.
**Abbreviations:** VCI, vascular cognitive impairment; NVCI, non-vascular cognitively impaired; SD, standard deviation; SE, standard error; ANCOVA, analysis of covariance; MoCA, Montreal Cognitive Assessment; VCAT, Visual Cognitive Assessment Test; WMH, White Matter Hyperintensities; DM, Diabetes Mellitus; HTN, Hypertension; HLD, Hyperlipidemia; VCS, vascular cognitive score

The highest average AUC for detecting VCI was observed with the CatBoost (AUC 0.85) and XGBoost (AUC 0.84) models, detailed in **Table 4**. Both models outperformed MoCA (AUC 0.65) and the Recognaize-VCS Composite score (AUC 0.77), with CatBoost also achieving the highest accuracy (0.85), sensitivity (0.85), and F1 score (0.82).

**Table 4.** Diagnostic performance of machine learning–based Vascular Cognitive Scores (VCS) and standard tools for detecting VCI.

| Score | Avg. AUC<br>(5-fold cv<br>for ML<br>models) | Avg.<br>Accuracy | Avg.<br>Sensitivity | Avg.<br>Specificity | F1<br>score |
| --- | --- | --- | --- | --- | --- |
| **MoCA** | 0.65 | 0.63 | 0.61 | 0.62 | 0.60 |
| **Recognaize-vcs-composite** | 0.75 | 0.73 | 0.73 | 0.74 | 0.70 |
| **Recognaize-vcs-catboost** | 0.85 | 0.80 | 0.85 | 0.84 | 0.82 |
| **Recognaize-vcs-xgboost** | 0.84 | 0.84 | 0.83 | 0.83 | 0.81 |
| **Recognaize-vcs-**<br>**randomforest** | 0.81 | 0.73 | 0.61 | 0.81 | 0.70 |
| **Recognaize-vcs-lightgbm** | 0.81 | 0.79 | 0.87 | 0.73 | 0.78 |

**Note.** CatBoost, XGBoost, Random Forest, and LightGBM are evaluated using 5-fold cross-validation. For each model, the average AUC, accuracy, sensitivity, specificity, and F1 score across the folds are reported.
**Abbreviations:** VCS, vascular cognitive scores

#### Novel digital cognitive features predictive of VCI

Feature importance ranking using SHAP values for the CatBoost model identified the most predictive features of VCI as *gs1_avg_success_time* [Grocery Shopping Average Success Time] (mean SHAP value +1.15) and *sm_time_iqr* [Symbol Matching IQR](+0.88), followed by age (+0.64), variability in round time, and total task completion time, as shown in Figure 4. These findings indicate that fine-grained, time-based behavioral metrics from cognitive game performance provided stronger discriminatory power than traditional cognitive test scores, especially in cerebrovascular cognitive impairments.

**Figure 4.**
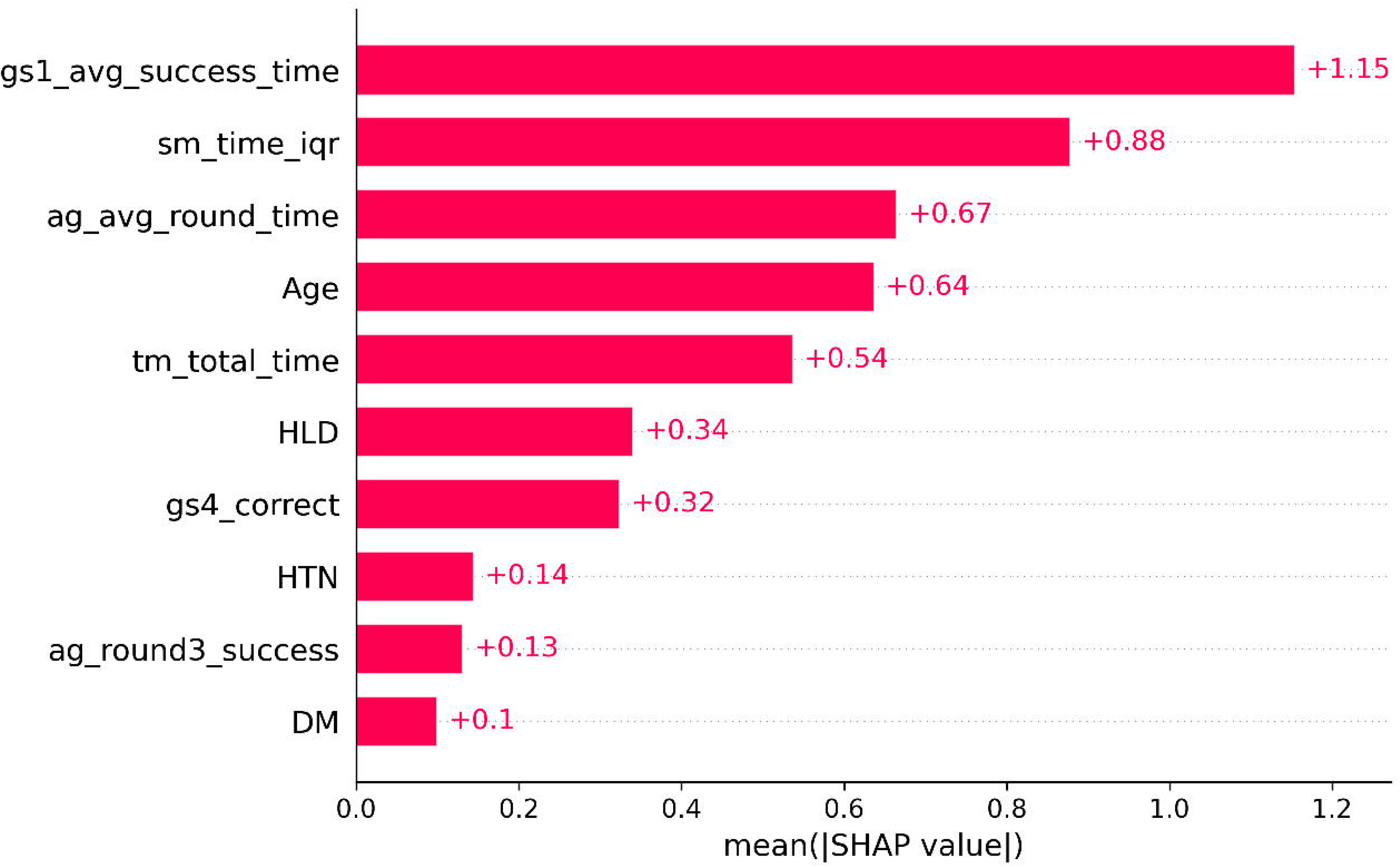
Feature importance ranking based on SHapley Additive exPlanations (SHAP) values for the CatBoost classification model used to distinguish VCI from NVCI. Higher mean absolute SHAP values indicate greater contribution to the model’s predictions. Key predictive features revealed novel digital biomarkers of VCI such as Grocery Shopping average success time (gs1_avg_success_time) and Symbol Matching response time variability (sm_time_iqr), which aligns with known clinical findings of executive dysfunction and reduced processing speed in cerebrovascular disease.

## 4. DISCUSSION

In this study, we developed and validated ReCOGnAIze, a gamified, AI-powered digital cognitive assessment tool designed to detect both MCI and VCI. Grounded in cognitive-behavioral science, the tool incorporates engaging tasks with automated scoring and interpretable ML models. We validated a *Recognaize_total* composite score which achieved an AUC of 0.9 for distinguishing MCI from CN, outperforming MoCA and VCAT, each of which achieved AUC less than 0.7. In addition to detecting MCI, we used explainable AI for identifying digital cognitive features for differentiating VCI from NVCI, revealing key insights about cerebrovascular-driven cognitive impairment. We then validated ML models for detecting VCI from NVCI, using ReCOGnAIze features and VCI risk variables, achieving an average AUC of 0.85. These findings suggest that ReCOGnAIze may serve as an effective and scalable cognitive screening tool, with interpretability and clinical relevance for identifying both MCI and VCI.

Compared to established tools such as MoCA, which reports AUCs ranging from 0.70 to 0.85 across diverse populations[64], ReCOGnAIze achieved comparable or higher AUCs while targeting cognitive domains underrepresented in traditional assessments. These include processing speed, response inhibition, and sustained attention—domains commonly impaired in VCI and non-amnestic MCI subtypes [18]. This task-specific evaluation might explain its high discriminative accuracy. The app’s brief duration (10–15 minutes), gamified design, and automated output enhance its potential for large-scale screening. While other digital tools have shown promise in detecting MCI, few have been validated specifically against neuroimaging or cerebrovascular biomarkers which show associations with non-amnestic domain impairments.

A key strength of ReCOGnAIze lies in its capability to detect VCI—an area largely overlooked by existing digital platforms despite the high global prevalence. Our explainable AI driven feature selection process highlighted novel digital biomarkers: response time variability, impulse control, and task switching—in line with executive impairments associated with VCI [6,18]. To the best of our knowledge, this is one of the first digital tools for the targeted detection of vascular-specific cognitive impairment using explainable feature attribution.

With explainable AI and clinical domain expertise, we prioritized model transparency and interpretability without compromising performance. After cross-validation, we used SHAP to generate individualized explanations of the predictions, facilitating clinician understanding and trust. While several studies have applied black-box models to cognitive classification, few have incorporated interpretable AI in a clinically meaningful way. Our pipeline ensures that each retained feature—such as response time variability, inhibition errors—has both a statistical and neuropsychological rationale, strengthening its relevance for clinical deployment in screening for MCI and VCI.

Key strengths of this study include the use of a deeply phenotyped, community based cohort with Brain-MRI, integration of cognitive-behavioural science in game design, and application of rigorous, multi-stage ML. Cognitive validity was established using a generalizable composite score, and ML models for differential detection were evaluated using cross-validation and out-of-fold predictions, reducing overfitting risk.

Nonetheless, several limitations must be acknowledged. First, the cross-sectional nature of the study precludes evaluation of predictive validity for future cognitive decline or dementia conversion. Longitudinal studies are needed to determine prognostic utility. Second, external validation in diverse, low-literacy, and rural populations is required. Third, cerebrovascular burden was assessed using WMH severity, which—while clinically relevant—does not capture all aspects of cerebrovascular pathology. Finally, performance in real-world clinical or community settings remains to be evaluated.

The findings of this study have several implications for clinical practice and public health. ReCOGnAIze offers a scalable, self-administered digital cognitive assessment that could be deployed in primary care or community settings for cognitive screening. Its short duration, engaging interface, and minimal training requirements make it feasible for wide-scale use. Importantly, the tool evaluates VCI—an under-recognized but potentially treatable contributor to dementia, with a high global burden.

## Conclusion

ReCOGnAIze is a novel, interpretable digital cognitive assessment that enables accurate detection of both MCI and VCI. By combining gamified tasks with explainable AI, it offers a transparent, efficient, and clinically relevant approach to early cognitive screening. This tool holds promise for transforming dementia risk stratification in both clinical and population health contexts, advancing precision medicine approaches to cognitive care.

## Data Availability

Selective data produced in the present study can be made available upon reasonable request to the authors and corresponding author.

## ACKNOWLEDGEMENTS, SOURCES OF FUNDING AND DISCLOSURES FUNDING SOURCES

This study received funding support from the Strategic Academic Initiative grant (SP1CLNT900-NTU-A630-PJ-03INP001400A630) from the Lee Kong Chian School of Medicine, Nanyang Technological University, Singapore, National Medical Research Council, Singapore under its Clinician Scientist Award (MOH-CSAINV18nov-0007), Ministry of Education Start-up Grant, Ministry of Education Academic Research Fund Tier 1 (RT02/21) and Ministry of Education Science of Learning grant (MOESOL2022-0002), NTUitive GAP Fund from Nanyang Technological University, Singapore (NGF-2023-13-019) and the Innovation to Startup Grant (I2Start-2307024) from the Singapore-MIT Alliance for Research and Technology (SMART) under the National Research Foundation (NRF), Singapore.

## AUTHOR CONTRIBUTIONS

AAM contributed to the conception, study design, data acquisition and analysis and drafting of the manuscript and figures.

AV contributed to the study design, data acquisition and drafting of the manuscript. YJL contributed to the data acquisition and drafting of the manuscript.

ES contributed to data acquisition and drafting of the manuscript and figures. FT contributed to data acquisition and drafting of the manuscript.

HA contributed to data acquisition and drafting of the manuscript. KXL contributed to data acquisition and drafting of the manuscript. PT contributed to data acquisition and drafting of the manuscript. GKS contributed to data acquisition and drafting of the manuscript. JDJW contributed to data acquisition and drafting of the manuscript. BQ contributed to data acquisition and drafting of the manuscript.

KA contributed to data acquisition and drafting of the manuscript.

NK contributed to conception, study design, data acquisition and analysis and drafting of the manuscript.

## DATA SHARING STATEMENT

Data used in this study will be shared upon reasonable request made to the Corresponding Author.

